# Early Identification of SARS-CoV-2 Emergence in the DoD via Retrospective Analysis of 2019-2020 Upper Respiratory Illness Samples

**DOI:** 10.1101/2021.02.18.21251368

**Authors:** Richard R. Chapleau, Monica Christian, Benjamin Connors, Christa Premo, Tim C. Chao, Juan Rodriguez, Shana Huntsberger, Jennifer Meyer, Amanda Javorina, Kenney Reynolds, David Riddle, Mark Lisanby, Clarise Starr

**Affiliations:** Applied Technology and Genomics Division, Public Health and Preventive Medicine Department, US Air Force School of Aerospace Medicine

## Abstract

The first case of non-travel related SARS-COV-2 was detected late February 2020 in California, however the delay of diagnostic testing and initial stringent testing criteria made it difficult to identify those who could have acquired it through community spread. There was speculation by many that the virus may have been circulating at least a month earlier [1], and environmental sampling has suggested that versions of this virus were found many months before the first human samples were identified [2]. Here we performed a retrospective study from residual samples collected from a global DoD Respiratory Surveillance Program to establish a tentative timeline by which this virus was circulating in our DoD population. We performed RT-PCR for SARS-COV-2 and compared to the dates of these cases to the first cases identified in respective states and counties using the Johns Hopkins COVID tracker website. Twenty-four positive samples were identified out of approximately 7,000 tested. Although we found some early cases in certain regions, we did not see circulation before late February in samples collected both in the US and outside the USA.

**SUMMARY:** *What is already known about this topic?:* We know that the first reported case of SARS-COV-2 was mid-January; however, there has been conjecture that the virus was found in the community before this date.

*What is added by this report?:* Here we took samples collection from a global respiratory surveillance program and evaluated for the presence of SARS-COV-2 RNA. The first case we found in the surveillance program was approximately 30-60 days before the first case was identified on military installations via diagnostic testing, however was not earlier than the mid-January reported case in California.

*What are the implications for public health practice?:* The implementation of new and emerging pathogen detection assays into already established surveillance programs could detect early community spread and possibly reduce spread of pathogen among vulnerable populations.

## METHODS

### Sample Selection

De-identified nasopharyngeal swab (NPS) samples collected from a respiratory surveillance program that tracks influenza globally to aid in the yearly vaccine development. Samples were stabilized in viral transport media prior to testing, covered the date range from 1 December 2019 to 03 June 2020, and represented 86 military medical treatment facilities (MTFs) around the globe. USAFSAM (a DoD Reference Laboratory) previously tested these samples for upper respiratory infections using the Luminex Respiratory panel (Austin, TX) before being stored at - 80ºC. Metadata provided by the laboratory and included date of collection, MTF, and co-infections.

RNA was extracted from the NPS samples with Promega Maxwell 16 instruments using the Maxwell Total Viral RNA kit according to the manufacturer’s instructions. We performed quantitative reverse-transcriptase PCR using the SuperScript III RT-PCR master mix (ThermoFisher, cat. #204454) with the research-use only 2019-nCoV primer-probe kit (IDT DNA, cat. #10006605). Thermocycling conditions on the ABI 7500 FAST analyzer was a 20-minute reverse transcriptase step at 50°C, a 10-minute hot-start activation step at 95°C, and 45 cycles of 95°C for 3 seconds followed by 55°C for 30 seconds.

### Data Analysis and Statistics

In accordance with the approved CDC assay methodology, samples were positive for SARS-CoV-2 if the RNase P control passed (Ct < 40) and both primer sets N1 and N2 produced Ct values below 40. In the cases where RNase P did not amplify but both N1 and N2 were positive, we called the sample positive in accordance with the CDC emergency use authorization guidance (revision 03, page 34). Samples where only one of the two markers were detected were “inconclusive” and the test was repeated using the previously extracted RNA. Metadata were stored in a separate computer from the analytical results and the Laboratory Director combined the PCR results with metadata prior to analysis. We used Microsoft Access for data management and descriptive statistics such as number of positives, daily positive hit rate, and earliest detection.

## RESULTS

Our study identified 24 positive samples from 7,021 total samples (0.3%). Of these samples, 14 were collected from patients prior to the first COVID-19 case clinically reported at their respective installations (Table 1). The first case identified at Wright Patterson AFB in Ohio was collected a full month before the first laboratory-confirmed case at that installation, and two days before the first laboratory-confirmed case in the Department of Defense (reported 26 February by US Forces Korea [Defense.gov, 2020]). We also identified a second case of significant interest 46 days before the first case reported from the installation in North Dakota. We received samples from installations in three geographic combatant commands (Table 2): European Command (EUCOM), Pacific Command (PACOM), and Northern Command (NORTHCOM). The majority of test samples (89.3%) originated in NORTHCOM followed by PACOM (6.6%), and EUCOM (3.4%). An additional 47 samples originated in US Coast Guard clinics (0.7%). We identified 20 positive samples from NORTHCOM (0.3% positive rate), zero samples from PACOM, four samples from EUCOM (1.7% positive rate), and zero positive samples from the Coast Guard.

**Table 1:**
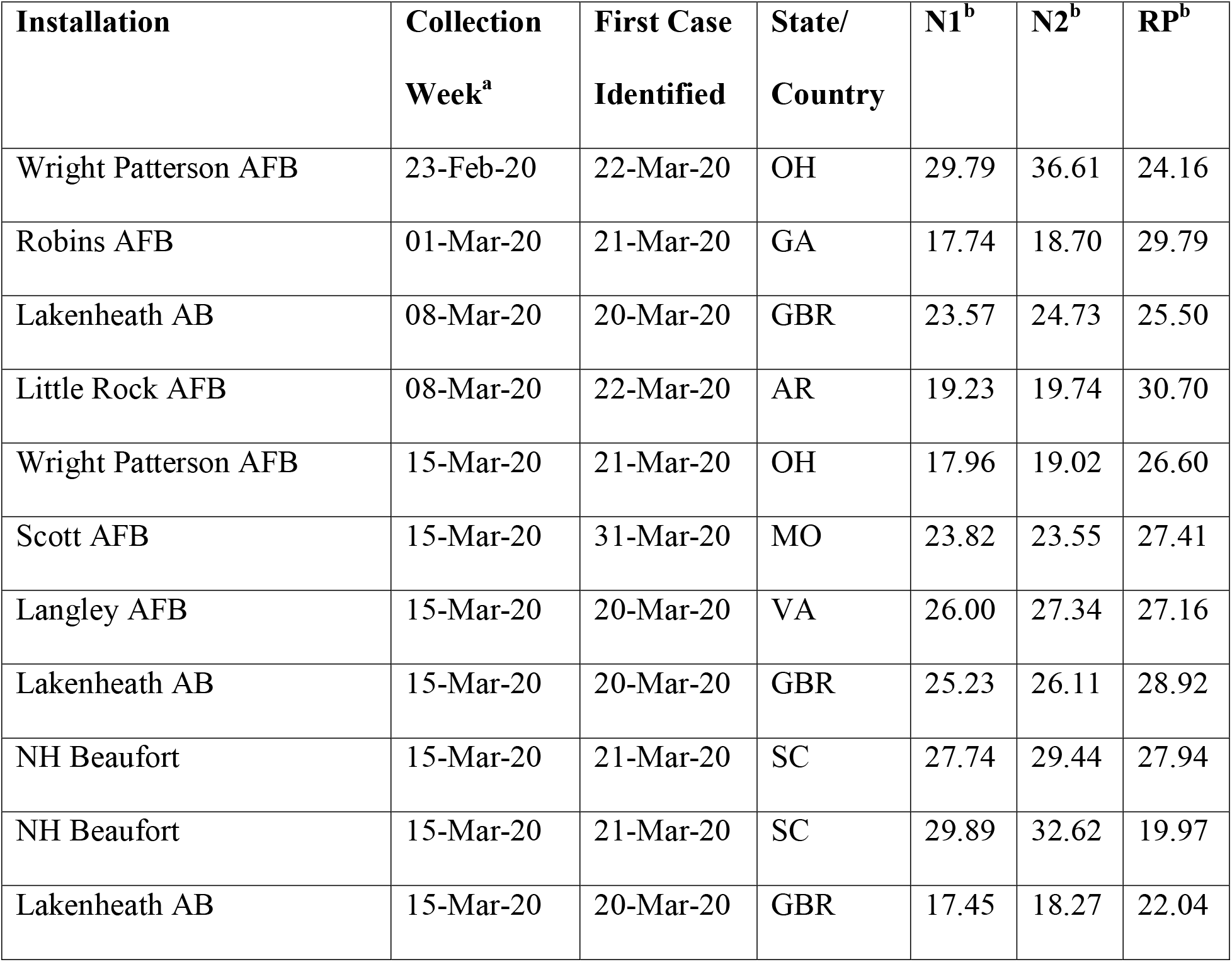
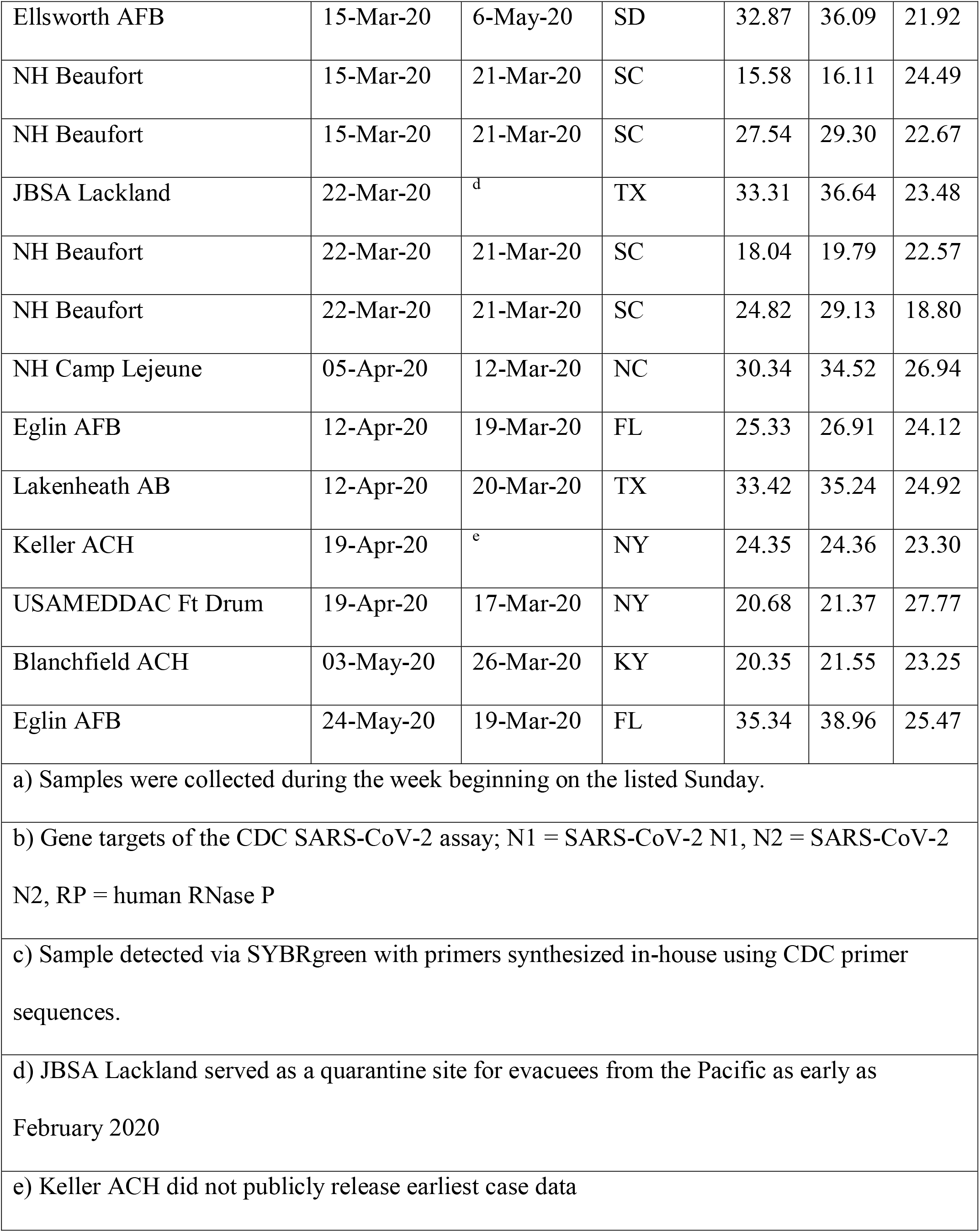
Summary table of SARS-CoV-2 positive samples.

**Table 2:**
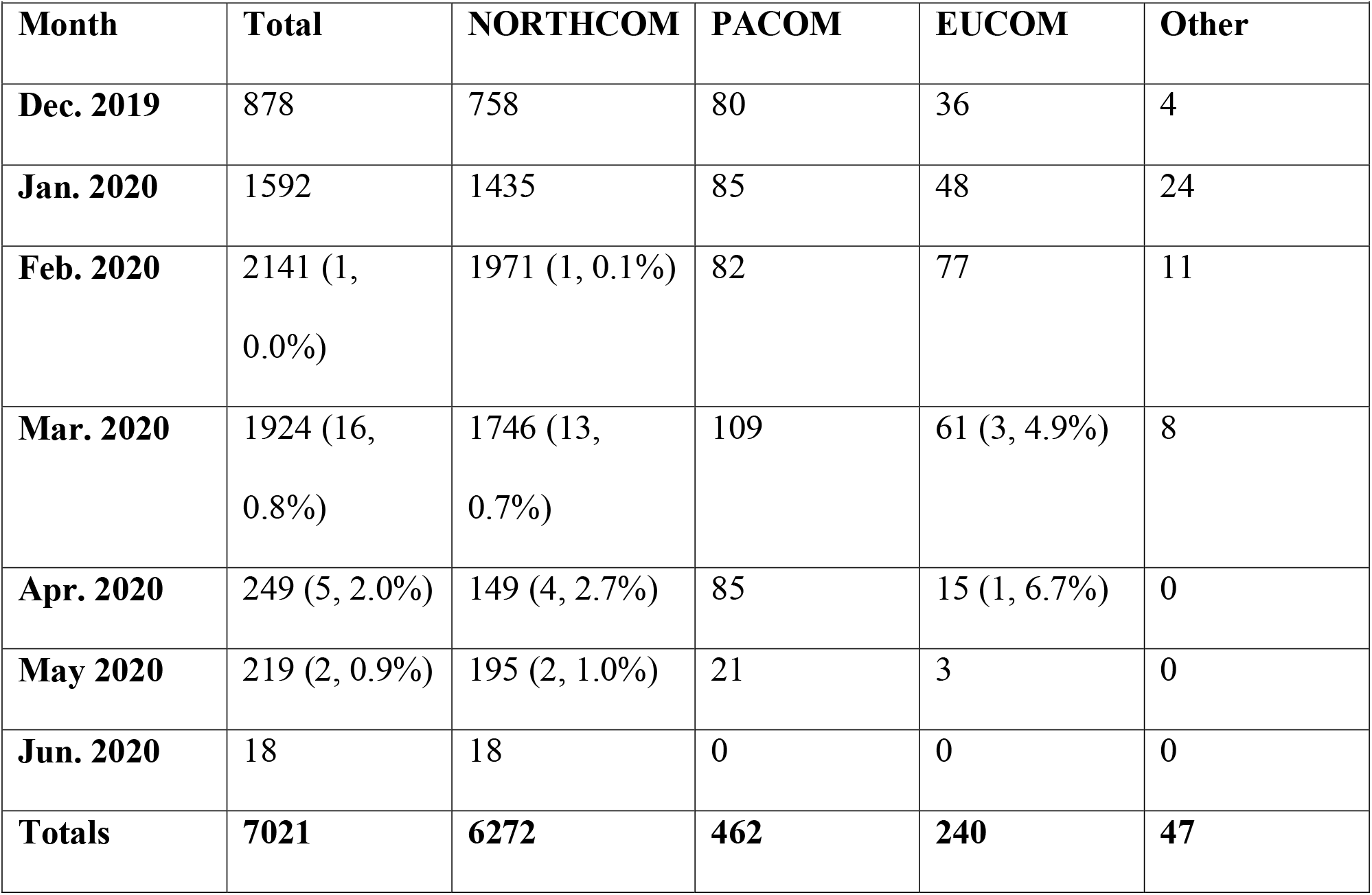
Samples tested by month and region.

| Table 2: Samples tested by month and region. |  |  |  |  |  |
| --- | --- | --- | --- | --- | --- |
| **Month** | **Total** | **NORTHCOM** | **PACOM** | **EUCOM** | **Other** |
| **Dec. 2019** | 878 | 758 | 80 | 36 | 4 |
| **Jan. 2020** | 1592 | 1435 | 85 | 48 | 24 |
| **Feb. 2020** | 2141 (1, 0.0%) | 1971 (1, 0.1%) | 82 | 77 | 11 |
| **Mar. 2020** | 1924 (16, 0.8%) | 1746 (13, 0.7%) | 109 | 61 (3, 4.9%) | 8 |
| **Apr. 2020** | 249 (5, 2.0%) | 149 (4, 2.7%) | 85 | 15 (1, 6.7%) | 0 |
| **May 2020** | 219 (2, 0.9%) | 195 (2, 1.0%) | 21 | 3 | 0 |
| **Jun. 2020** | 18 | 18 | 0 | 0 | 0 |
| **Totals** | **7021** | **6272** | **462** | **240** | **47** |

As expected, our paradigm of testing patient residual samples remaining from other molecular testing for upper respiratory resulted in detecting a logarithmically increasing positivity rate (Figure 1). Beginning the week ending 29 February, we observed an increasing number of positive tests until the week ending 28 March. The decline at the end of March was due to the addition of SARS-CoV-2 clinical testing in the Air Force clinical test menu in early March. Therefore, most of the SARS-CoV-2 positive samples in March and beyond were evaluated directly by the clinical lab and not by the surveillance lab. The effect of clinical testing was also seen in that the peak sampling period was from mid-February to early March, when nearly 1/3 of all samples were collected. Notably, the effect was most dramatically seen in testing numbers between the weeks ending 21 March and 28 March, where we tested 691 samples compared with 90, respectively.

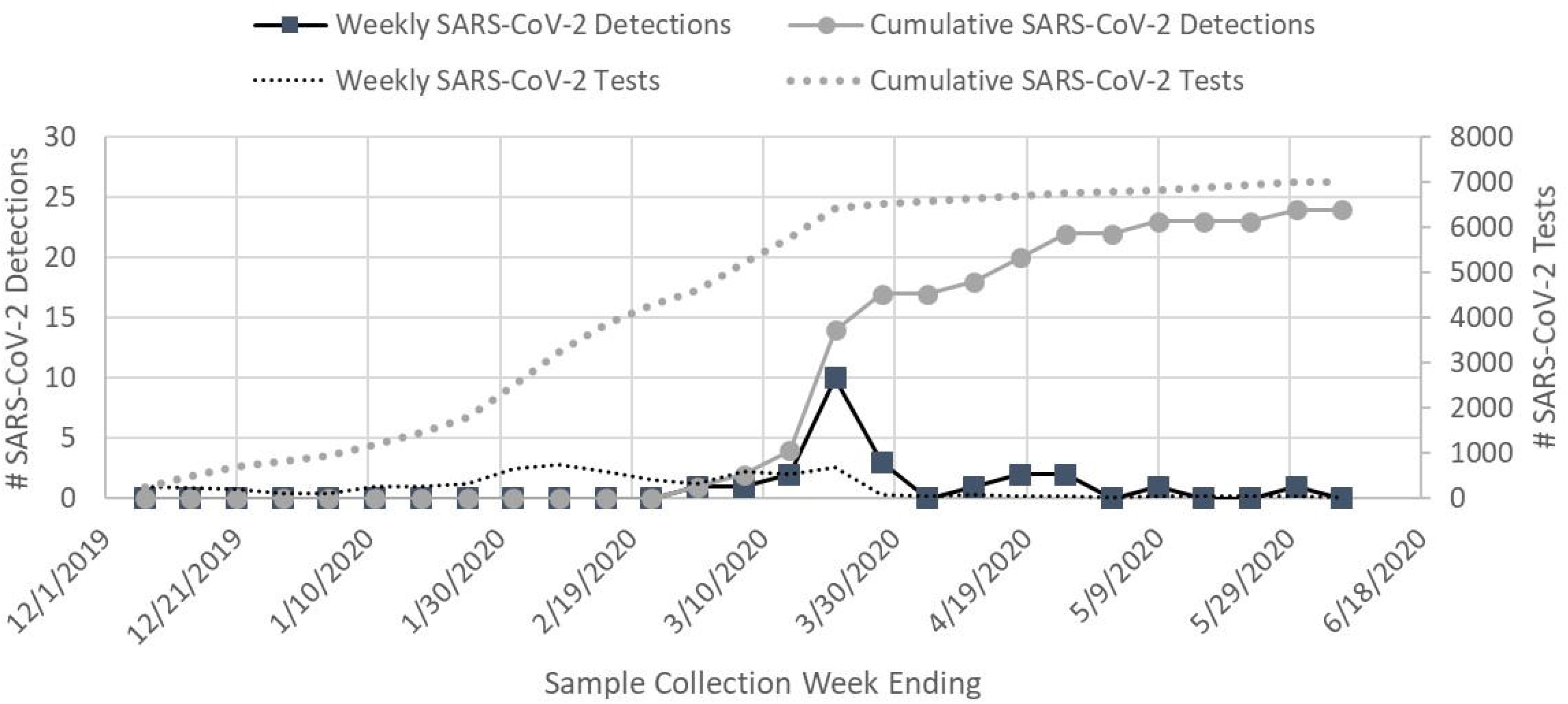

In addition to observing an increase in testing, we detected two peaks the positivity rates. For the week ending 29 February, we detected one positive sample of 333 tests, a rate of 0.30%. The following two weeks remained below 1% positive rate (0.17% and 0.37%, respectively) before increasing to 1.45% with 10 detections of 688 tests for the week ending 21 March, and peaking at 3.33% with three positive tests of 90 samples during the final week of our study. The week ending 28 March had no positive tests, but then the next three weeks had positivity rates of 1.4%, 3.1%, and 3.8% before another week of no tests and finally a week with 2.0% positivity (1/49 tests). These observations are consistent with the observed increase in nationwide infection growth beginning during the week ending 14 March [3].

## DISCUSSION

Our results suggest that while the epidemic was present in the US military population earlier than previously reported, the change is minimal. Excluding the outlier from Ellsworth AFB, we identified initial positive cases approximately two weeks prior to the first case reported (average = 11.6 days, range 2 to 27 days prior). Our results are similar to other retrospective testing that focused on PCR-based detection of residual samples [4-6]. Evaluation of residual specimens using antibody tests is still an on-going effort and may lead to different data as better assays comes available [7].

Due to early resource limitations in the global molecular testing supply chain, we began testing our early samples using the SYBRgreen detection method. Several of the samples ran out of material prior to establishing consistent supply chain provisions for probe-based detection using the CDC assay and Superscript real-time detection. One of these samples was the detected sample from late February.

While we tested more than 7,000 samples, the number of independent test sites is small and heavily favored by one Service. We expected a high proportion of samples to originate from NORTHCOM facilities, as most of the DoD MTFs are located there, however representation overseas is limited with only ten Air Force installations and one Army installation. In a demonstration of collaboration, we did receive samples from six Coast Guard clinics, too. Additionally, we did not receive any samples from Southern Command and less than 10 samples from Central Command, so the impact of the epidemic in our military members stationed in those regions is unknown. In order to provide adequate surveillance during normal operations and epidemics, the Military Health System, and the Defense Health Agency by extension, must increase the participation of sentinel sites in the Respiratory Surveillance Program. With 475 military hospitals and medical clinics across the globe, the DoD is the only healthcare system that is equipped to monitor worldwide infectious diseases before they enter the homeland. The surveillance network for the DoD is primed to immediately add validated assays for new and emerging infections, before the clinical assays are deployed, to actively track community spread once a virus has been detected by travel or any criteria that are tracked early on in a pandemic by the CDC.

## Data Availability

Summary data of threshhold crossover cycles are available from the corresponding author upon reasonable request.

